# EXAMINING THE DOUBLE BURDEN OF MALNUTRITION FOR PRESCHOOL CHILDREN AND WOMEN OF REPRODUCTIVE AGE IN LOW- AND MIDDLE-INCOME COUNTRIES: A SCOPING REVIEW PROTOCOL

**DOI:** 10.1101/2021.06.17.21259113

**Authors:** Jason Mulimba Were, Saverio Stranges, Ishor Sharma, Juan-Camilo Vargas-González, M. Karen Campbell

**Author notes:** **Correspondence to:** Jason Mulimba Were, PhD Candidate, Department of Epidemiology and Biostatistics, Schulich School of Medicine & Dentistry, Western University, Kresge Building Room 201, London, Ontario, Canada, N6A 5C1.

## Abstract

**Introduction:** The majority of the populations in Low-and Middle-Income Countries (LMICs) are encountering the double burden of malnutrition (DBM): the coexistence of both undernutrition and overnutrition sequalae. With DBM being a new phenomenon in research, little is known about its etiology, operational definitions and risk factors influencing its manifestation. The proposed scoping review is aimed at mapping literature with regards to the DBM phenomenon among preschool children and women of reproductive age in LMICs who are among the most high-risk groups to encounter DBM.

**Methods:** A comprehensive literature search will be conducted in the following electronic databases: MEDLINE, EMBASE, Scopus, CINAHL, LILACS and ProQuest Dissertations & Thesis Global. Additionally, searches in other government and institutional sources (World Health Organization website and university repositories) and forward and backward citation tracking of seminal articles will also be done. Two reviewers will independently conduct title and abstract screening and full text screening. Similarly, data extraction and coding will independently be done by two reviewers. Information extracted from included literature will be analysed qualitatively using thematic analysis approach and reported as per the Preferred Reporting Items for Systematic reviews and Meta-Analyses extension for Scoping Reviews (PRISMA-ScR) guidelines.

**Ethics and Dissemination:** Ethical approval is not required for this study because the review is based on literature from publicly available sources. The dissemination of our findings will be done through presentations in relevant conferences and publication in a peer-reviewed journal.

**Strengths and limitations:**

- To the best of our knowledge, this is the first scoping review that focuses on exploring the etiology of the double burden of malnutrition among preschool children and women of reproductive age in Low-and Middle-Income Countries.
- This review will encompass comprehensive literature search and will utilize a renowned thematic analysis framework to synthesis the findings of the study.
- The findings of this review will be important in not only mapping the current literature with regards to the double burden of malnutrition phenomenon for risk populations but also guiding secondary data analysis for our subsequent studies.
- The anticipated dearth of causation literature and longitudinal studies in this area of research may limit our findings, specifically in understanding the etiology of double burden of malnutrition.

## INTRODUCTION

The modern-day world is characterised by the paradoxical co-occurrence of nutrition-related diseases and conditions. On one hand, overnutrition is fast becoming a global public health problem. Recent data from the World Health Organization (WHO) indicates that approximately 2 billion adults (above 18 years) and 40 million preschool children (under 5 years of age) are overweight or obese.[1] This is a likely reflection of the ongoing nutrition transition, primarily in Low-and Middle-Income Countries (LMICs), that is currently characterized by the progressive affinity for calorie-dense foods and sedentary lifestyles in many population subgroups.[2,3] On the other hand, nutritional deficiencies that are facilitated by food inadequacies still persistent in many population subgroups around the world, namely in the Global South.[2] Nearly half a billion adults have been estimated to be underweight while 155 million preschool children have been reported to be stunted.[1] This contemporaneous existence of multiple nutrition burdens has led to the emergence of the double burden of malnutrition (DBM), a construct that characterizes the simultaneous presence of both undernutrition and overnutrition sequalae.[1] The DBM can be manifested at three different levels[1]: individual level - depicting the co-occurrence of stunting, wasting or micronutrient deficiency with overweight/obesity at a single point in time or across the life course; household level - where at least two household members have discordant nutritional statuses; and the population level - where both undernutrition and overnutrition types are common in the same population unit.

Although everyone is susceptible to changes in nutritional health in response to socio-demographic changes, women of reproductive age (WRA) and preschool children are among the most vulnerable population groups.[3,4] These groups’ risks of DBM are partly independent of each other due to differences in their physiological states (i.e., reproductive stage for women and critical development stage for preschool children) that are highly sensitive to nutritional changes, and partly dependent due to the close contact between the preschool children and their mothers (dependency of the preschool children on the mother for nourishment and care).[5,6] Furthermore, some of the proximate risk factors of DBM among these two groups are by themselves influenced by broader socio-demographic factors acting at different levels of the society (e.g., household, community, and regional levels).[5]

The emergence of DBM creates an opportunity for health practitioners to identify population sub-groups at most risk of adverse health outcomes. In recognition of this, the WHO in 2016 developed the *double-duty actions*, to provide a framework that leverages existing policies, interventions, and programs to concurrently reduce the risk of DBM.[1] However, there is a dearth of studies from LMICs to spur the actualization of this and other related frameworks. A major contributor to this scarcity is the recency of the concept of DBM in research. The definition of DBM as the coexistence of under- and overnutrition within individual’s, households, population, or life-course is broad enough to capture the current nutrition realties in a lot of the world’s population.[6,7] However, this broad definition does not offer clear guidance on the specifics of measuring DBM. In extant literature, DBM has been defined in a myriad of ways, using varied nutritional biomarkers.[6,7] Presumably, the thinking underlying these definitions have been based on study context, target population and data availability.[7,8] Further, there is no clear understanding as to whether the co-occurrence of the malnutrition indicators in the documented literature represents a non-random cluster with plausible similar etiology. Therefore, this study intends to conduct a scoping review with the aim of examining the rationale underlying the malnutrition phenotypes used in DBM studies focusing on preschool children and WRA.

## METHODS AND ANALYSIS

A scoping review was determined as the most suitable literature synthesis design for this study given the exploratory nature of the study objective.[9] After gaining familiarity with the current literature on DBM among the populations of interest in low-resourced setting, we established the following research objective: To describe the current state of knowledge regarding the double burden of malnutrition among preschool children and women of reproductive age in low- and middle-income countries. This objective will be examined through the following research questions:

1. Which nutrition indicators have been used to operationally define the double burden of malnutrition among preschool children and women of reproductive age?
2. What are the posited explanations for the occurrence of the identified double burden of malnutrition phenotypes among preschool children and women of reproductive age?
3. What are the risk factors for the double burden of malnutrition among preschool children and women of reproductive age?

Although there are systematic reviews (both published and ongoing reviews) that have focused on the operational definitions of the DBM,[7,10,11] to the best of our knowledge, we found no existing reviews that succinctly synthesises literature on DBM phenomenon based on the review questions that we have formulated.

### Patient and public involvement

There will be no patient and public involvement in this review.

### Eligibility criteria

This study will use the Population-Concept-Context (PCC) framework in determining the study’s eligibility criteria.[9] The populations of interest will be women of reproductive age (15-49 years) and children below the age of five years (0-59 months). The key concept is the double burden of malnutrition defined as the coexistence of undernutrition and overnutrition burdens. Studies will be included if they contain any of the following key concepts: Measurement of DBM at the individual, household, population, or life-course levels; theoretical frameworks, models or body of knowledge used to explain DBM phenotypes; and risk factors for DBM. The context includes all studies that were conducted in LMICs-as defined by the world bank.[12]

There are no limitations with regards to the study design. Peer-reviewed articles, gray literature, theses, dissertations, published and unpublished reports will be included. Abstracts for which primary data or full text literature can be accessed will also be included. All non-English studies will be included. In instances where studies are published in native languages, a native speaking person will be requested to translate the study into English. Furthermore, no limits will be placed with regards to the time-period of which the study was conducted. Studies that solely focus on men, children above the age of five years, adults above 50 years and LMIC migrants residing in High-Income-Countries (HIC) will be excluded. Further, studies will be limited to those involving human participants.

### Search strategy

Discussions with a library specialist and among co-authors identified relevant databases. A collective decision was to search the following electronic databases: MEDLINE, EMBASE, Scopus, CINAHL, LILACS and ProQuest Dissertations & Thesis Global. The search will be conducted using a combination of the subject headings and keywords relating to ‘double burden of malnutrition’, ‘women of reproductive age’, ‘preschool children’ and ‘low- and middle-income countries. Additionally, relevant gray literature from government sources and organizations such as the WHO will also be searched. After screening, forward, and backward citation tracking of seminal studies will be done to identify all relevant articles missed in the database search. Authors of unpublished data will be contacted to provide any relevant information. After completing the searches, the reference list will be uploaded in EndNote citation management software[13] for de-duplication and afterwards transferred to Rayyan software[14] for screening purposes.

### Screening

Screening will be done according to the Preferred Reporting Items for Systematic reviews and Meta-Analyses extension for Scoping Reviews (PRISMA-ScR) guidelines.[9] Prior to the review screening, all reviewers will conduct a pilot screening of 10 randomly selected articles and discuss their findings to achieve consistency in the application of the eligibility criteria during the screening process.[15] Afterwards, there will be two levels of article screening: Level 1 will entail screening of study titles and abstract to eliminate obvious irrelevant articles.[9] Level 2 will constitute full-text screening for all eligible studies against the inclusion/exclusion criteria. Both levels will be done by two reviewers independently.[9] Disagreements will be resolved through discussions with a third reviewer. The PRISMA diagram will be used to highlight the outcome of the screening process (Figure 1).

**Figure 1:**
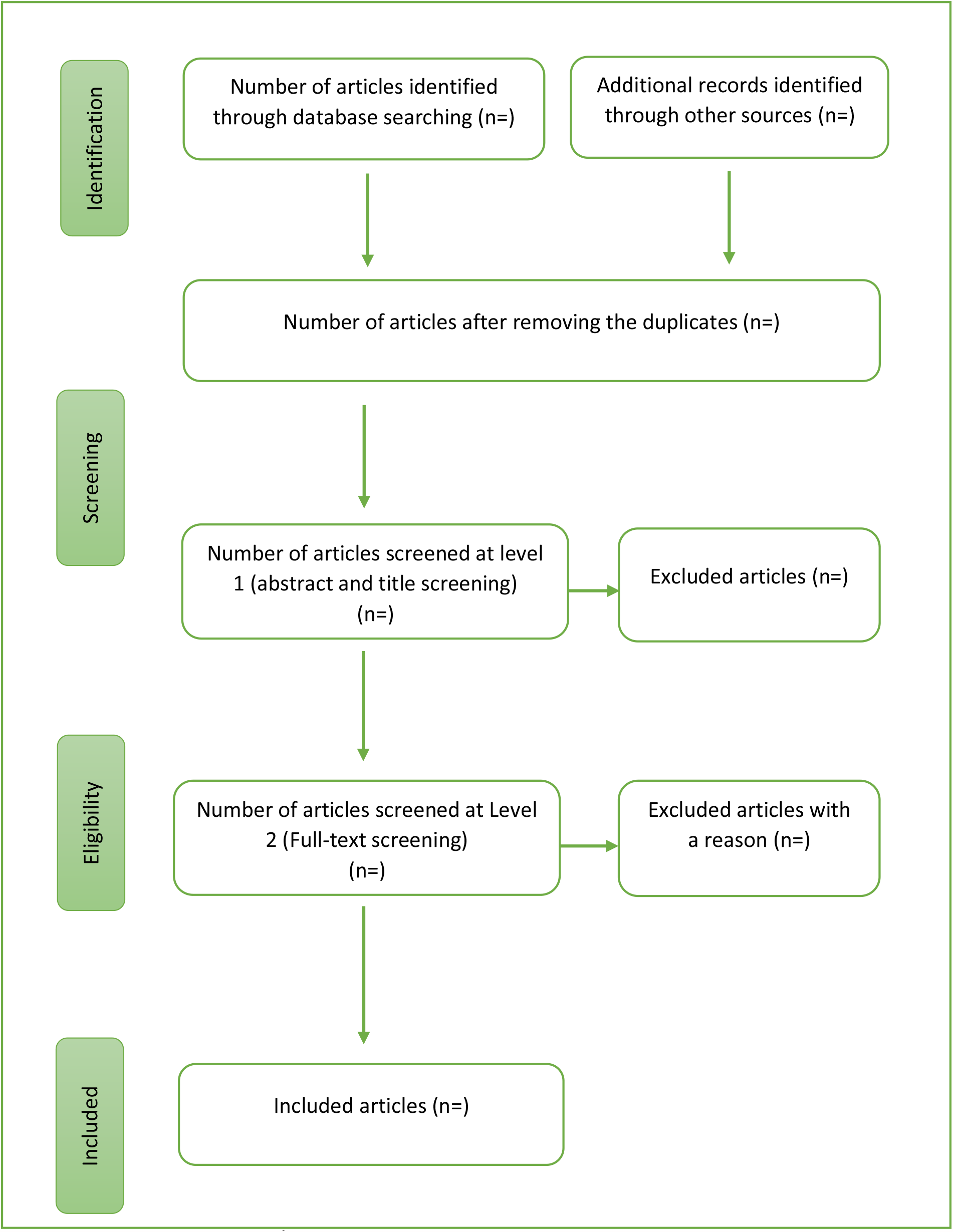
Scoping Review Flow Diagram.

### Data charting process

Two reviewers will independently extract data from the selected articles using a charting form developed by the review team. In general, the form will include bibliographic information, study characteristics (context, study design, and participants characteristics), concept (malnutrition indicators, DBM definitions, level of DBM analysis, theoretical/conceptual frameworks), significant findings and conclusions (Table 1). To ensure consistency and accuracy in recording data while using the charting form, a pilot test will be done by the reviewers independently on 10% of the included studies.[15] The charting form will be calibrated based on the feedback from the pilot study.[9] Afterwards, the review team will continuously update the charting form in an iterative manner to ensure all relevant information is captured during the charting process. Any disagreements will be resolved through discussion with a third reviewer.

**Table 1:** Sample Data Charting Form.

| Category | Description |
| --- | --- |
| Author/Year | Author(s) name(s) and year of publication |
| Study Title | Working title of the study |
| Aim/Objectives | A description of the aim of the study as stated by the authors |
| Setting/Context | Country where the study was conducted. |
|  | Specific Location in which the study was conducted (district/province/institution based). |
|  | Year the study was conducted. |
| Methods | Study design used to conduct the study (e.g., cohort study, case control, cross-sectional) |
|  | Participant's characteristics (Specify salient characteristics of the target population e.g., sex and age bracket) |
|  | Data sources used in the study (primary or secondary-specify the source) |
|  | Sampling strategy: Briefly specify how participants were identified and recruited the study |
|  | Data collection: Specify how data was gathered (e.g., measurement tools, questionnaires etc.) |
|  | Outcome measures: Specify malnutrition indicators that have been used to measure nutritional status and how the double burden of malnutrition was operationally defined |
|  | Exposure measures: specify the main covariates that were studied |
|  | Data analysis: Specify the analytical strategy (e.g., thematic analysis, logistic regression etc.) |
| Results | Operational definition of DBM: Describe all the findings with regards DBM phenotypes that have been used (i.e., nutritional biomarkers used in the definition and reported prevalence) |
|  | Explanations for the occurrence of DBM: Extract any information that provides explanations/hypothesis for the cooccurrence of the reported DBM phenotypes |
|  | Exposure: Discuss any reporting of factors (risk or protective factors) found to influence the occurrence of DBM |
|  | Miscellaneous findings: Report any other important findings |
| Limitations | Highlight the shortcomings of the study |

### Data analysis and synthesis

This study will present a qualitative synthesis of current literature performed using the inductive thematic analysis as suggested by Braun & Clarke (2008).[16] Extracted information will be subjected to content analysis, structured based on the review objectives as follows: study context, population type, malnutrition indicators, DBM phenotypes, explanations for the occurrence of DBM phenotypes and risk factors for DBM. Each reviewer will independently re-read the extracted information and afterwards, generate codes and apply them to the excerpts of extracted information.[16] All the codes and related data excerpts will be collated and sorted into potential themes.[16] The resulting themes will then be reviewed and refined by the review team with the aim of examining the relationship between the themes and the review questions.[16] Finally, a narrative synthesis and visual display (where possible) will be performed for the generated themes.[16]

### Conclusion

The proposed scoping review will be conducted as first phase of a study the seeks to understand the DBM phenomenon in sub-Saharan Africa. We believe that the information gathered in this review will provide an up-to-date understanding of DBM phenomenon in LMICs. Consequently, synthesis of DBM knowledge will provide a foundation for assessing the current operational definitions and identifying important knowledge gaps in literature. Furthermore, findings from the proposed scoping review will guide future analytical research including a secondary data analysis for the second phase of this study. It is anticipated that the proposed review will inform the modeling and interpretation of DBM outcomes for our subsequent studies. Additionally, the proposed review is expected to provide more insights on the potential risk factors of DBM and the interrelationships among them which will be used to guide the development of statistical models (path diagrams) that would be empirically tested in the second phase of the study.

## Data Availability

This is a scoping review protocol that intends to map data from publicly available sources. No data has been gathered.

## ETHICS AND DISSEMINATION

Ethical approval is not required for this study because the review is based on literature from publicly available sources. The dissemination of our findings will be done through conference presentations and publication in a peer-reviewed journal.

## Acknowledgements

We would like to express our gratitude to Piotr Wilk and Shehzad Ali (Faculty members, Department of Epidemiology and Biostatistics at Western University) for their advisory support for this project. We would also like to thank Marisa Tippet (Research and Scholarly Communications librarian at Western university) for her technical guidance in the development of the study’s methods section.

## Authors’ Contribution

JMW, SS, and MKC decided on the area of research. JMW, SS, IS, JCVG and MKC conceptualized the research, contributed to the development of the review questions and writing of the manuscript. SS and MKC were responsible for supervising the project. JMW was responsible for the final editing, formatting, and submission.

## Patient consent for publication

Not required.

